# Does BCG protect against SARS-CoV-2 infection ?: elements of proof

**DOI:** 10.1101/2020.05.01.20087437

**Authors:** Yassine Ouanes, Mokhtar Bibi, Nesrine Baradai, Marouane Boukhris, Kays Chaker, Aziz Kacem, Houcem Hedhli, Kheireddine Mrad Deli, Ahmed Sellami, Sami Ben Rhouma, Yassine Nouira

## Abstract

**Background:** There are several factors explaining the difference in the spread of SARS-CoV-2 infection including the BCG vaccination. This fact is supported by the concept of beneficial non specific effect of this live vaccine associated to its interaction with the immune system.

Our study aims to identify the correlation between the universal BCG vaccination policy and the mortality attributed to COVID-19.

**Methods:** We conducted an epidemiological study in which we collected COVID-19 pandemic data of April 11^th^, 2020 from the web site worldometers.info. The exclusion criteria for our study were a number of inhabitants less than one million, low-income countries according to the World Bank classification, a total number of infection cases less than 500 and countries that have performed less than one hundred tests per million inhabitants.

**Results:** Countries that never had universal BCG vaccination policy have a higher mortality (correlated to performed diagnostic tests) attributed to SARS-CoV-2 infection (p<0.001).

We found that the year of introduction of vaccination influenced significantly the mortality. Countries that started immunization policy before 1960 had more favorable results (p=0.049).

For countries that started the BCG vaccination after 1960, countries with current policies have lower mortality attributed to SARS-CoV-2 infection than countries that have stopped immunization (p=0.047).

**Conclusions:** Countries that have a BCG vaccination policy have a lower mortality attributed to SARS-CoV-2 infection. The populations of countries that applied this immunization before 1960 are more protected even if this universal policy has been interrupted.

## INTRODUCTION

Covid-19 has spread around the world, however, its impact varies from one country to another. Indeed, there is a significant difference even between comparable countries in terms of health system and containment measures.

This finding is certainly attributed to a multitude of intricate factors including the variability of individual immune system reaction to this infection. Recent epidemiological studies have developed the concept of beneficial non specific effect of live vaccines. This means that it may have effects on infections not targeted by the vaccine.^1^

Observationl studies found that Bacillus Calmette Guerin (BCG) vaccination is associated with reduced non specific mortality and better survival^1^. Other immunological studies demonstrated that BCG can induce trained innate immuniy. Otherwise, urologists know well that BCG is an attenuated mycobacterium developed as a vaccine for tuberculosis that has demonstrated antitumor activity in several different cancers including bladder cancer.^2^

These concordant elements, in favor of the non specific immunotherapeutic mechanism of the BCG vaccine, led us to make the assumption that the COVID-19 mortality can be partially explained by the vaccination policies of countries around the world.

Based on these observations, we hypothesized that countries which have an early start of universal BCG vaccination policy would have a reduced morbidity and mortality attributed to SARS-CoV-2 infection.

## METHODS

We collected data concerning the global BCG vaccination policies and practices across countries from the second edition of the BCG World Atlas.^3^ COVID-19 pandemic informations of April 11^th^, 2020 were obtained from the web site worldometers.info.

The mortality may be influenced by a country’s standards of the health system. For this reason, we classified countries using the gross national income per capita in 2018 according to the Worls Bank data.^4^

The exclusion criteria of our study were the following: a number of inhabitants less than one million, low-income countries according to the World Bank classification, a total number of infection cases less than 500 and countries that have performed less than one hundred tests per million inhabitants.

We classified the countries according to the World Bank classification into 4 groups because we think that the mortality may be influenced by the standard of medical care. Group (a) includes lower middle income countries that have universal BCG vaccination policy. Group (b) includes upper middle and high income countries that have a current universal BCG vaccination policy. Group (c) includes upper middle and high income countries that had universal BCG vaccination policy but which has been stopped. Group (d) includes countries that never had universal BCG vaccination policy.

For a better comparison between countries we have defined the index Rd which is equal to deaths attributed to SARS-CoV-2 infection per million inhabitants (D) divided by the number of diagnostic tests per million inhabitants (T) multiplied by 1000 which gives the following mathematical formula Rd = D / T x 1000.

Two groups were identified according to their index Rd: high risk group with a Rd index over 1.95 (median of Rd) and low risk group with a Rd index below this value.

To study the evolution of mortality over time we applied a Kaplan-Meier survival analysis. It was used to measure the fraction of patients living for a certain amount of time after applying the BCG vaccination therapy.

## RESULTS

Initially, we compared the number of deaths per million inhabitants attributed to SARS-CoV-2 infection (D) in the four groups of countries.

Upper middle and high income countries that never had universal BCG vaccination policy (the group d includes United States, Italy, Netherlands, Belgium and Lebanon) have a significant higher mortality rate (mean mortality rate = 148.8 deaths / million) than the author groups of countries (p<0.001). (Fig. 1)

**Figure 1:**
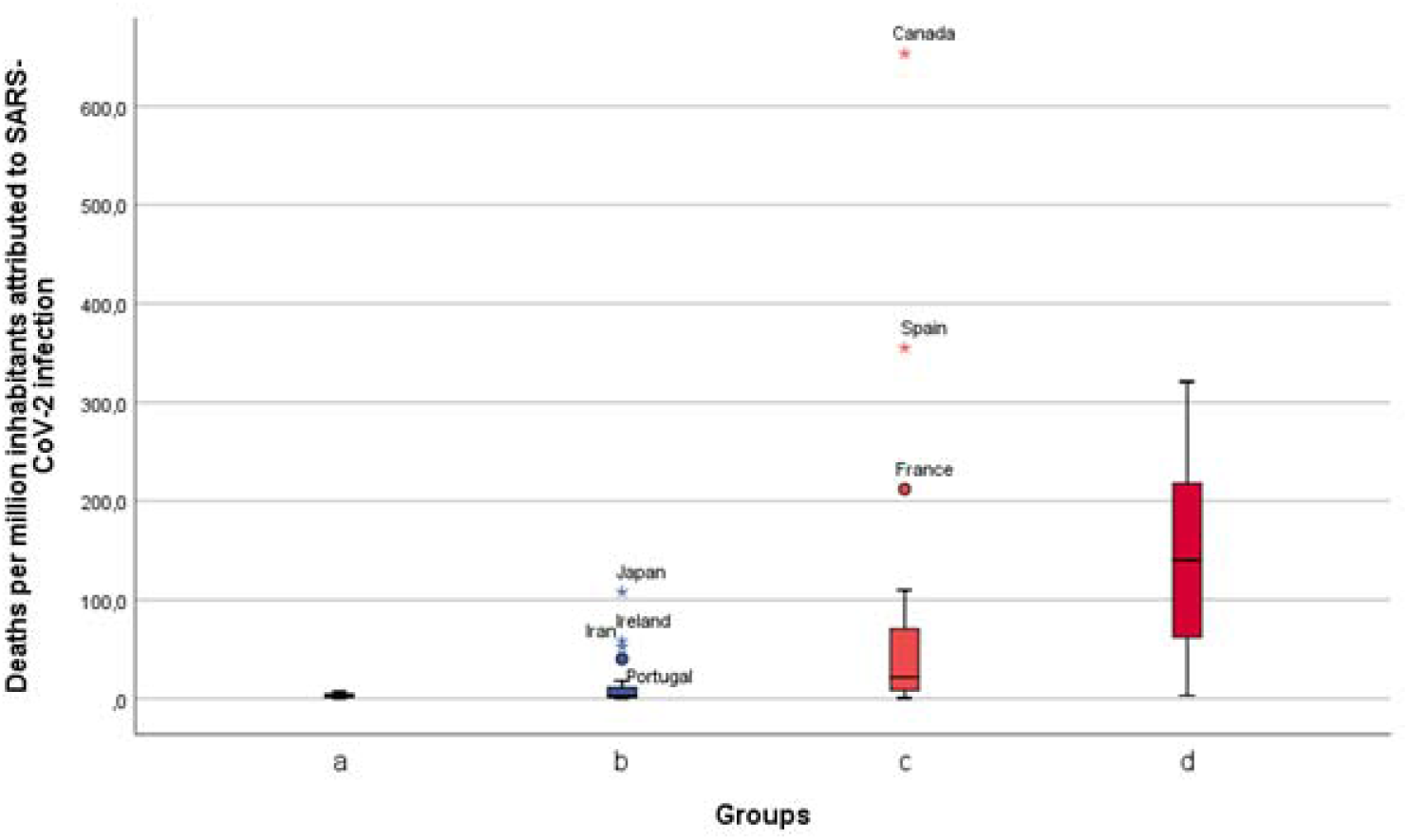
Mortality attributed to SARS-CoV-2 infection in the four groups of countries.

Given the great variability between countries in terms of diagnostic tests performed (T = number of performed tests per million inhabitants), we calculated the index Rd for a better analysis (Rd = D / T x 1000).

Upper middle and high income countries that never had universal BCG vaccination policy have a significant higher Rd index (19.21 for group d Vs 2.47 for group a, 1.5 for group b and 1.87 for group c; p<0.001). (Fig. 2)

**Figure 2:**
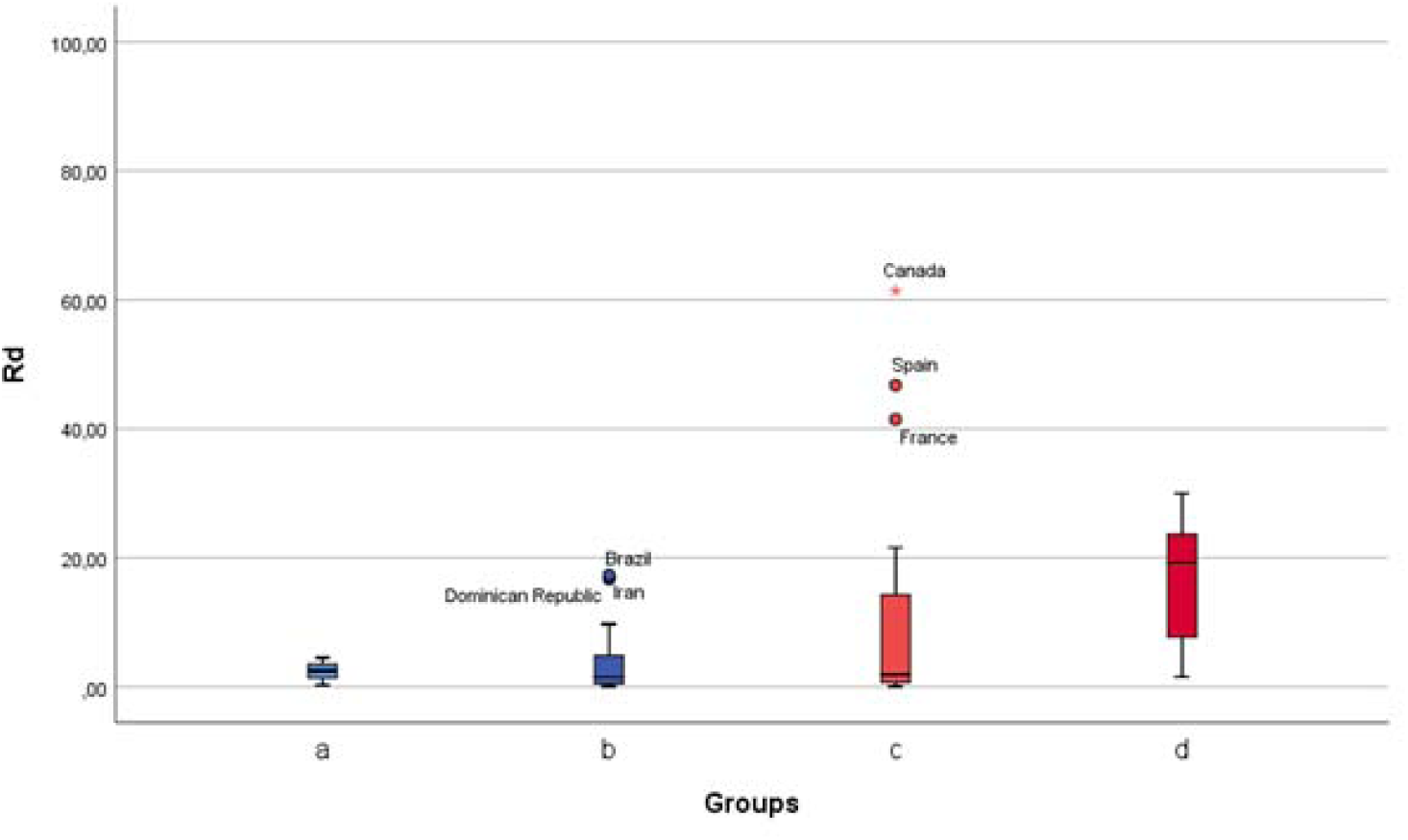
Mortality Correlated to diagnostic tests (index Rd) attributed to SARS-CoV-2 infection in the four groups.

At this level of our study, we concluded that countries which applied a universal BCG vaccination policy had a lower mortality attributed to COVID-19. This fact led us to wonder if the duration of this vaccination policy was a determining factor given that advanced age is a risk factor for SARS-CoV-2 infection.

According to the Figure 3, countries which have a current BCG vaccination policy that lasted for at least 60 years had better results.

**Figure 3:**
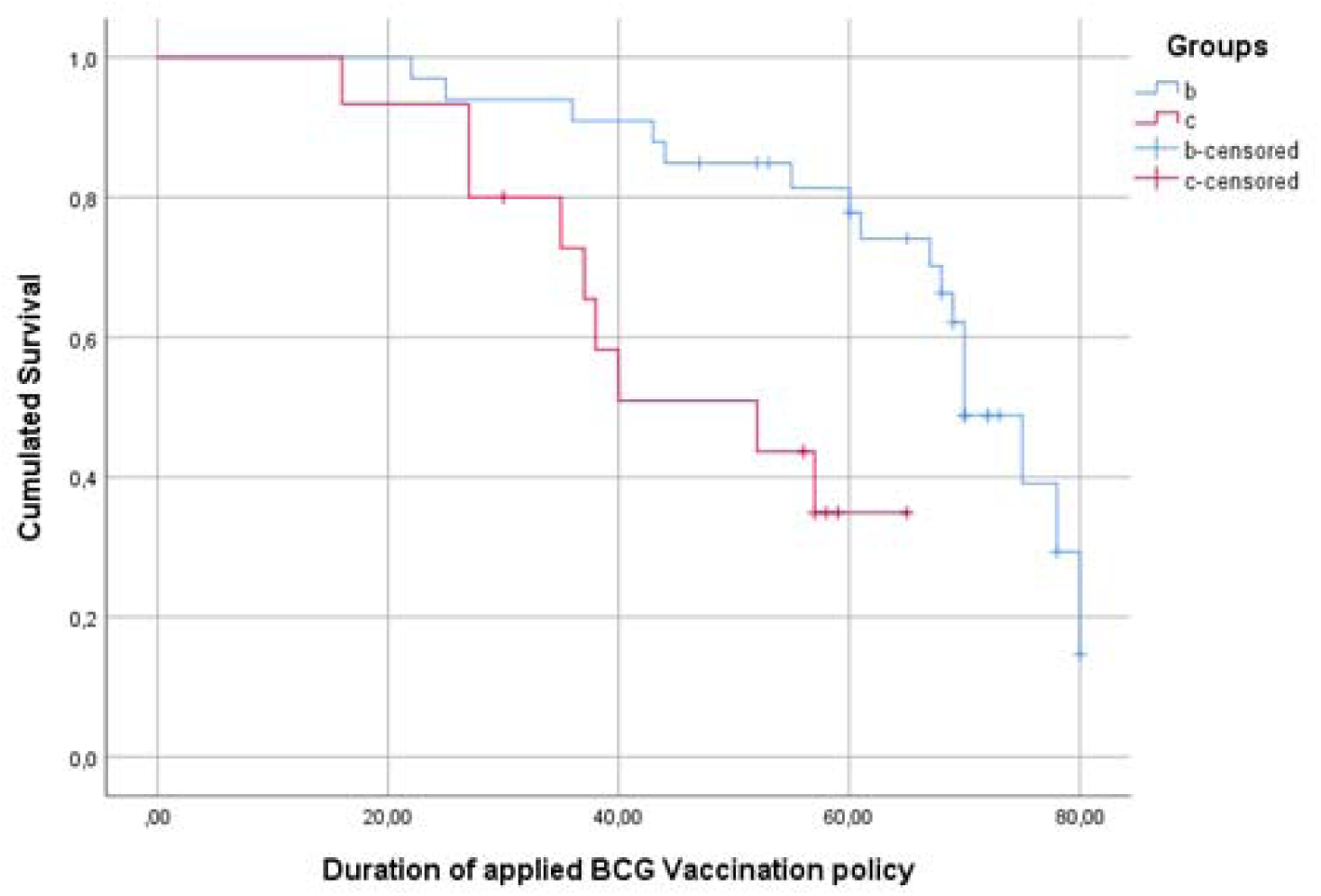
Analysis of the cumulated survival in upper middle and high income countries which had a universal BCG vaccination policy.

In the next part of our study, we analyzed the effect of BCG according to the year of introduction of vccination. As shown in Figure 4, it seems that the older this vaccination, the better. After 1960, upper middle and high income countries that have a current universal BCG vaccination policy (group b) have a better cumulated survival than upper middle and high income countries that had universal BCG vaccination policy but which has been stopped (group c).

**Figure 4:**
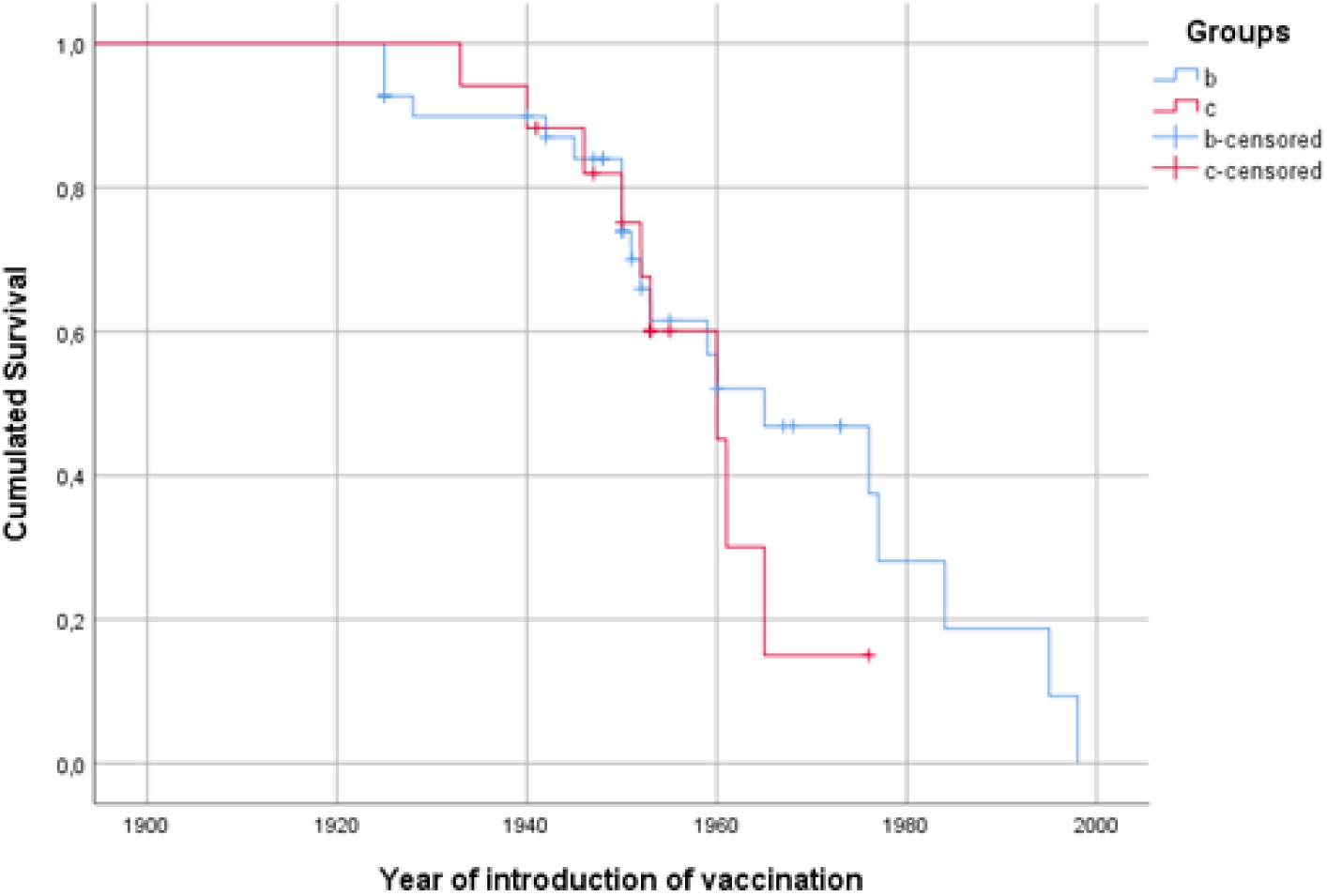
The impact of the year of BCG vaccination introduction in the upper middle and high income countries which had an immunization policy.

The Kaplan-Meier graph showed that the year 1960 was a critical year that influenced the survival within the different studied countries. ROC curve analysis showed the same result as the threshold is year 1960 (AUC=0.647[0.509-0.785]; p=0.049).

In the last part of our study, we compared the countries whose populations were vaccinated with BCG relatively to this threshold year (1960). It should be noted that all the countries in the group a applied a vaccination policy before 1960. (Fig. 5)

**Figure 5:**
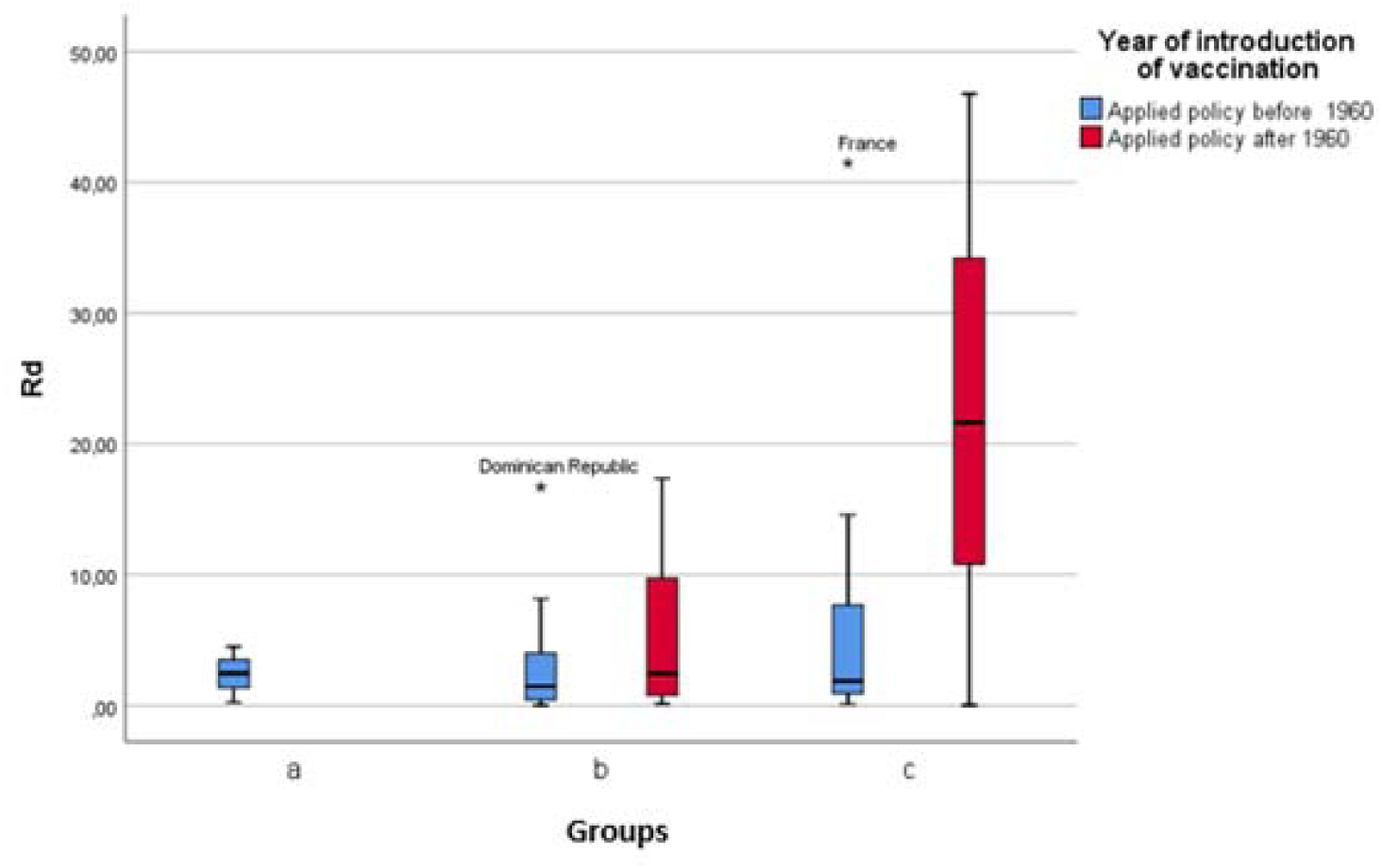
Comparison of countries which applied a BCG vaccination policy before or after 1960.

For upper middle and high income countries that have a current universal BCG vaccination policy (group b), the index Rd does not differ according to the application of the vaccination policy before or after 1960 (p=0.41).

For upper middle and high income countries that had universal BCG vaccination policy but which has been stopped (group c), countries which applied the vaccination policy before 1960 have a more favorable Rd index (Rd=10.15 before 1960 Vs Rd=22.8 after). However, this result is not significant (p=0.676).

For countries that started BCG vaccination before 1960, there is no significant difference between countries that have an ongoing policy and those that stopped vaccination (p=0.901).

For countries which started the BCG vaccination after 1960, countries with current policies have a better Rd index than countries that have stopped immunization (Group b: Rd=5.58; Group c: Rd=22.8; p=0.047).

## DISCUSSION

The different impact of SARS-CoV-2 infection from one country to another is certainly due to a combination of factors. The situation in Italy is quite edifying with high morbidity and mortality, while it seems under control in other countries with a similar economic and health systems.^5^

Furthermore, containment measures have been taken all over the world every few days. We think that what differentiates a group of individuals from another is the immune system’s response to this pathogen. Hence the importance of vaccination to deal with infections. It has been thought that the role of a vaccine was to protect from a specific disease. However, numerous immunological studies have concluded that a vaccine can have a non specific protective effect.^1^

At the beginning of the 20^th^ century, Calmette already noted that children vaccinated with BCG had a lower mortality rate^1^. Since then may have been born the principle of non specific effects of BCG vaccine. BCG was used in the 1970s in clinical studies as a non specific immunotherapy for multiple malignant neoplasms. While severeal reports have show its efficiency in the treatments of melanoma and leukaemia, others were controversial making BCG use limited in such settings. Nonethless, BCG has continued to be used for non-muscle-invasive bladder cancer since its first use in 1976.^6,7^ Indeed, we currently know the mechanism of action of intravesical instillations of BCG and it seems that it is attributed to a local immune response characterized by induced expression of cytokines in the urine and bladder wall and by an influx of granulocytes and mononuclear and dendritic cells.^8^

Faced with this set of elements, we hypothesized that BCG vaccination could have a protective effect against SARS-CoV-2 infection.

Initially, we found that upper middle and high income countries that never had universal BCG vaccination policy (group d) have a significant higher mortality rate.

In order to reduce the bias due to diagnostic tests, we applied the Rd index which is equal to deaths attributed to SARS-CoV-2 infection per million inhabitants (D) divided by the number of diagnostic tests per million inhabitants (T) multiplied by 1000 (Rd = D / T x 1000). So we found the same observation as the first graph with a significant higher Rd in middle and high income countries that never had universal BCG vaccination policy (group d). We concluded that BCG vaccination was a protective factor against SARS-CoV-2 infection.

Recent studies concerning COVID-19 showed that advanced age represents an independent risk factor for this infection.^9,10^ For this reason, we tried to establish whether the age of vaccination was a protective factor, especially for the older adults. We hypothesized that the older the vaccination, the better. Therefore, we investigated the impact of the year of BCG vaccination introduction in each country.

We concluded that upper middle and high income countries that have a current universal BCG vaccination policy (group b) have a better cumulated survival than upper middle and high income countries that had universal BCG vaccination policy but which has been stopped (group c) after 1960. We noticed that countries which applied a universal BCG vaccination policy before 1960 are protected against SARS-CoV-2 infection even if immunization ceased a few decades later. On the other hand, countries which applied a late vaccination policy (after 1960) and continue immunization are better protected than those that have stopped it. We therefore concluded that 1960 was a pivotal year.

We conducted a comparative analysis which confirmed these findings. Regarding the countries that started BCG vaccination before 1960, there was no significant difference between countries that have an ongoing vaccination policy and those that stopped it (p=0.901) few decades later. Whereas countries which started the BCG vaccination after 1960 and those with current policies have a better Rd index than countries that have stopped immunization (Group b: Rd=5.58; Group c: Rd=22.8; p=0.047).

Our explanation is that the older adults are the main target of the SARS-CoV-2 infection and BCG vaccination is therefore a protective factor for the population currently aged 60 or over. Indeed, this finding corroborates the results of other studies which have concluded that the efficacy of BCG is generally maintained up to 60 years after vaccination carried out during childhood.^11^

Our study provided some elemnts of proof that universal BCG vaccination interfers favorably with COVID-19. However, a randomized controlled trial is required to confirm this finding and determine the necessary duration that the immune system needs to develop protection against SARS-CoV-2 infection.

## CONCLUSIONS

Or results revealed that countries that never had universal BCG vaccination policy have a higher mortality (correlated to performed diagnostic tests) attributed to SARS-CoV-2 infection. We also highlighted that BCG vaccination was a protective factor against this disease.

We noticed that countries which applied a universal BCG vaccination policy before 1960 are better protected against SARS-CoV-2 infection even if immunization ceased a few decades later. On the other hand, countries which applied a late vaccination policy (after 1960) and continue immunization are better protected than those that have stopped it.

## Data Availability

The datasets generated during and/or analysed during the current study are available from the corresponding author.

## REFERENCES

1. Aaby P, Benn CS. Developing the concept of beneficial non specific effect of live vaccines with epidemiological studies. Clin Microbiol Infect. 2019;25:1459–67.

2. Morales A, Eidinger D, Bruce AW. Intracavitary bacillus Calmette Guerin in the treatment of superficial bladder tumors. J Urol. 1976;116:180–3.

3. Zwerling A, Behr M, Varma A. 2011. The BCG World Atlas: a database of global BCG vaccination policies and practices. PLoS Med. 8:e1001012. http://dx.doi.org/10.1371/journal.pmed.1001012.

4. World Bank Group, World Health Organization. Global Civil Registration and Vital Statistics: Scaling up Investment Plan 2015-2024. Washington, D.C.: World Bank Group; 2014. https://openknowledge.worldbank.org/bitstream/handle/10986/18962/883510WP0CRVS000Box385194B00PUBLIC0.pdf?sequence=1&isAllowed=y. Accessed 9 Aug 2019.

5. Mizumoto K, Kagaya K, Zarebski A. Estimating the asymptomatic proportion of coronavirus disease 2019 (COVID-19) cases on board the Diamond Princess cruise ship, Yokohama, Japan, 2020. Euro Surveill. 2020;25:2000180.

6. Van Der Meijden AP. Non-specific immunotherapy with B.C.G. in superficial bladder cancer: an overview. In Vivo. 1991;5:599–604.

7. Moorlag S, Arts R, van Crevel R. Non-specific effects of BCG vaccine on viral infections. Clin Microbiol Infect. 2019;25:1473–8.

8. Shen Z, Shen T, Wientjes MG. Intravesical treatments of bladder cancer: review. Pharm Res. 2008;25:1500–10.

9. Chen R, Liang W, Jiang M. Risk factors of fatal outcome in hospitalized subjects with coronavirus disease 2019 from a nationwide analysis in China. Chest. 2020;15:30710–18.

10. Wu JT, Leung K, Bushman M. Estimating clinical severity of COVID-19 from the transmission dynamics in Wuhan, China. Nat Med. 2020;26:506–10.

11. Aronson NE, Santhosham M, Comstock GW. Long-term efficacy of BCG vaccine in Americans Indians and Alaska natives. JAMA. 2004;291:2086–91.

